# Amyloid Screening and Treatment Drop Off in Patients with Left Ventricular Hypertrophy: Associations with Patient Socio-demographic Characteristics

**DOI:** 10.1101/2023.05.23.23290432

**Authors:** Kristie M. Walenczyk, Avinainder Singh, Kimhouy Tong, Matthew M. Burg, Edward J. Miller

**Affiliations:** Section of Cardiovascular Medicine, Department of Internal Medicine, Yale School of Medicine, New Haven, CT; VA Connecticut Healthcare System, West Haven, CT; Section of Cardiovascular Medicine, Brigham and Women’s Hospital, Boston, MA; Clinical Pharmacy Services, Yale New Haven Hospital, New Haven, CT; Department of Anesthesiology, Yale School of Medicine, New Haven, CT

## Abstract

**Background:** Compared to estimated population prevalence rates, relatively few patients at risk are ultimately diagnosed with and treated for transthyretin cardiac amyloidosis (ATTR-CA). Where along the clinical imaging and treatment pathway patient drop off occurs, and the association of drop off at each step with patient socio-demographic characteristics remains unknown.

**Methods:** Using data from a healthcare system-wide cardiovascular imaging repository and specialty pharmacy we characterized the multi-level clinical pathway from screening for left ventricular hypertrophy (LVH) with transthoracic echocardiograms (TTE), diagnosis with technetium-99m pyrophosphate scintigraphy (PYP), and tafamidis prescription, initiation, and adherence. Standardized differences (*d*≥0.20 indicating a small effect size or larger) were used to compare socio-demographics (age, sex, race, national and state Area Deprivation Index) among patients with TTE-identified LVH by PYP referral status, patients with PYP-identified ATTR-CA by tafamidis prescription status, and patients prescribed tafamidis by initiation status.

**Results:** A total of 8575 patients had LVH on TTE, with 1.6% referred for PYP. Of 97 patients with PYP-identified ATTR-CA, 58.8% were prescribed tafamidis, with 80.7% of those initiating therapy. Referral from LVH on TTE to PYP was higher among men, older patients, and those of Black/African American or American Indian/Alaskan Native race (all *d*’s≥0.20). Patients with PYP-identified ATTR-CA prescribed tafamidis were younger than those not prescribed tafamidis (*d*=-0.30). Utilization of a specialty pharmacy resulted in enrichment of treatment in subgroups traditionally undertreated in cardiovascular medicine, with higher rates of tafamidis initiation among women (100% initiation), patients of Black/African American race (*d*=0.40), and those living in more economically disadvantaged areas (*d*’s≥0.30).

**Conclusion:** These findings highlight the tremendous opportunity for more robust imaging and clinical ATTR-CA screening programs, including potential patient subgroups that should be targeted to reduce disparities. Among patients diagnosed with ATTR-CA, utilization of a specialty pharmacy process appears to ensure the equitable provision of tafamidis therapy.

## Introduction

Transthyretin cardiac amyloidosis (ATTR-CA) is a progressive and often fatal disease caused by misfolding and extracellular deposition of mutated or wild-type monomers of transthyretin that form amyloid fibrils (1,2). ATTR-CA can cause cardiac conduction abnormalities, arrythmias, and heart failure with preserved ejection fraction (HFpEF) (3-6), as well as systemic symptoms such as neuropathy (peripheral and/or autonomic), spinal stenosis, and biceps tendon rupture (7-9). Compared to estimated population prevalence rates, relatively few patients at risk are ultimately diagnosed with and treated for ATTR-CA (10). In many cases, the diagnosis is made late in the progress of the disease in the context of overt cardiac symptoms, which may be years after the initial symptom onset (3,10). However, with the introduction of targeted treatments for ATTR-CA such as tafamidis, there is a need for earlier identification of the condition. By preventing or slowing further amyloid deposition, tafamidis reduces the risk of hospitalization and mortality (11) and can potentially double a patient’s projected quality-adjusted life-years (12,13).

A transthoracic echocardiogram (TTE) is among the first diagnostic tests that can raise suspicion for ATTR-CA. TTE detection of classic features for ATTR-CA (such as concentric left ventricular hypertrophy (LVH)) may signal the need for further screening 3,10,14). Ideally in patients with clinical ‘red flag’ risk factors for ATTR-CA, LVH in the absence of other appreciable causes should prompt referral for diagnostic testing for ATTR-CA using established and accurate non-invasive diagnostic techniques such as technetium-99m pyrophosphate scintigraphy (PYP) (15). Subsequently, ATTR-CA positive patients should be offered initiation of transthyretin stabilizer therapy. In practice though, it remains unclear as to how often this clinical pathway is followed, and factors that account for patient drop off at each step from LVH identification on TTE, to ATTR-CA diagnostic testing with PYP scintigraphy, and ultimately to therapy initiation.

Among the factors that may play a role in the potential drop off from screening to treatment are patient socio-demographic characteristics. The prevalence of wild-type ATTR-CA is higher among men and increases with age (1,10), and the most common hereditary ATTR-CA variant in the United States (V142I) occurs almost exclusively among Black individuals with West African ancestry (16). Provider knowledge of this risk for ATTR-CA should result in a greater proportion of men, elderly, and Black patients being referred for PYP after detection of LVH. However, we know that Black patients, as well as other demographic groups (e.g., women and those of lower socioeconomic status) are less likely to undergo cardiac diagnostic testing (17-21) or to receive appropriate medical therapy (17,22-24). It is possible that these same disparities are also present in the screening, diagnosis, and treatment of ATTR-CA.

The cost of tafamidis remains extraordinarily high (25-27), and it may also be that patient socioeconomic status is associated with a provider’s decision to prescribe this therapy to an ATTR-CA positive patient, and whether a patient is subsequently able to start the medication. Due to the high cost and benefits authorization concerns, tafamidis is prescribed through a Specialty Pharmacy process. Specialty pharmacies serve to provide medications for conditions requiring high-cost complex therapies, monitoring, and/or special communication with patients, and provide services such as benefits verification, access to patient financial assistance programs, and assistance with prior authorization. While there is initial evidence from Medicare and specialty pharmacy databases suggesting that adherence to tafamidis transthyretin stabilizer therapy is good if it is prescribed (28,29), there is no data describing where in the clinical pathway the drop off of patient groups occurs and whether drop off was due to any identifiable patient characteristics.

Understanding the association of socio-demographic characteristics with the progression of patients from TTE screening for LVH, to PYP imaging diagnosis, and finally to tafamidis prescription and initiation can be used to inform provider and system-level efforts aimed at improving the screening, diagnosis, and treatment of ATTR-CA. We therefore used data from a single, diverse healthcare system – including a cardiovascular imaging repository and specialty pharmacy records – to determine the associations of patient age, sex, race, and neighborhood socioeconomic disadvantage with 1) PYP imaging among patients with TTE-detected LVH, and 2) tafamidis prescription, initiation, and adherence among patients diagnosed with ATTR-CA.

## Methods

### Study Sample and Design

This was a retrospective analysis of data from Yale-New Haven Health System (YNHHS), a large healthcare system in the Northeast United States. A health system-wide comprehensive cardiovascular imaging repository was queried to identify patients who underwent a TTE over a two-year period following the FDA-approval of tafamidis. We included patients aged 18 and older with LVH on TTE. We excluded patients: 1) missing data for interventricular septum (IVSd) or left ventricular posterior wall (LVPWd) thickness or 2) address, and 3) those with addresses outside of the typical 5 state catchment area (CT, MA, RI, NJ, and NY) of the healthcare system as their validity could not be verified.

We then reviewed the YNHHS cardiovascular imaging repository to determine which patients with LVH on TTE also underwent subsequent PYP imaging and that was positive for cardiac ATTR-CA. The time frame for PYP imaging was extended three months past that of the TTE search to allow time for test scheduling and completion post-TTE. The ATTR-CA diagnostic criteria of Gillmore et al. (30) was followed, including the exclusion of evidence of a plasma cell dyscrasia. We additionally queried the repository to identify all other patients with ATTR-CA-positive PYP imaging during this time to ensure capture of patients without TTE prior to PYP, TTE outside of our period of observation or completed at an outside institution, and those without LVH on TTE. For this combined sample representing all patients in the healthcare system with cardiac ATTR-CA confirmed via PYP, we then obtained tafamidis fill records from the YNHHS Specialty Pharmacy, which provides initial tracking of all tafamidis prescriptions written by YNHHS-affiliated providers, even if the prescription was ultimately dispensed by an external Specialty Pharmacy. This study was approved by the Yale University Institutional Review Board as exempt given that it was limited to retrospective review of medical record data collected during routine clinical practice.

### Study Variables

#### Socio-demographics

The following data were obtained from the electronic medical record for all patients – age at the time of TTE (or PYP for patients without LVH on preceding TTE), sex, race (self-identified), and address. Addresses were used to determine the patient’s census block group (derived from the nine-digit zip code) and corresponding Area Deprivation Index (ADI). The ADI is a validated and widely used measure of neighborhood socioeconomic disadvantage freely accessible through the University of Wisconsin Madison Neighborhood Atlas (31,32). The score is calculated based on 17 census-derived metrics related to resident average income, education, family composition, and housing quality (31,32). We recorded both national (range: 1 to 100) and state-level (range: 1 to 10) ADI rank, with higher scores indicative of greater disadvantage.

#### Imaging Data

Identification of LVH on TTE was defined as IVSd and/or LVPWd thickness of ≥13mm on TTE. Identification of ATTR-CA on PYP was based on established interpretive standards (33) as documented by the reviewing physician in the imaging report.

#### Pharmacy Data

For all patients with ATTR-CA confirmed via PYP, the YNHHS Specialty Pharmacy database was queried to determine – 1) prescription of tafamidis therapy and corresponding dates, and 2) initiation of tafamidis therapy defined as ≥1 tafamidis prescription dispensed to the patient. For patients with >1 dispensed tafamidis prescription, we determined the proportion of days covered (PDC), a measure of medication adherence. The PDC is calculated as a percentage of the medication days’ supply dispensed to the number of days between fills, with values >80% representative of a high adherence (34).

### Statistical Analysis

Cohen’s d/standardized difference (continuous variables) and standardized differences for proportions (categorical and dichotomous variables) were used to compare patient socio-demographics (i.e., age, sex, race, state ADI, and national ADI) among – 1) patients with TTE-identified LVH that completed PYP imaging vs. those without subsequent PYP imaging, 2) patients with PYP-identified ATTR-CA prescribed tafamidis vs. those not prescribed tafamidis, and 3) patients prescribed tafamidis that initiated therapy vs. those never filling the prescription. For these comparisons, a *d* of 0.20, 0.50, and 0.80 (absolute value) indicated a small, medium, and large effect size, respectively (35).

Among patients with >1 fill of tafamidis, the distribution of the PDC (medication adherence) was evaluated with the Kolmogorov-Smirnov test. The association of the PDC to patient socio-demographics was then evaluated with Pearson correlations (continuous variables) and the non-parametric Wilcoxon ranked sums (sex) and Kruskal-Wallis (race) tests. Analyses were performed using SAS software (v.9.4, SAS Institute, Cary, North Carolina, USA) and p<.05 (two-tailed) was used to determine statistical significance.

## Results

The patient identification flow is detailed in the consort diagram in **Figure 1**. A total of 8575 unique patients had LVH on TTE, of which 8461 (98.7%) met sample eligibility criteria. Among patients with LVH, 1.6% (n=138) were referred for PYP imaging with 42.7% (n=59) being diagnosed with ATTR-CA. Query of the PYP imaging data revealed another 38 patients with ATTR-CA without preceding TTE during the study period. Of the 97 total patients with PYP-identified ATTR-CA in the healthcare system, 58.8% (n=57) were prescribed tafamidis, with 80.7% (n=46) of those prescribed tafamidis initiating therapy.

**Figure 1.**
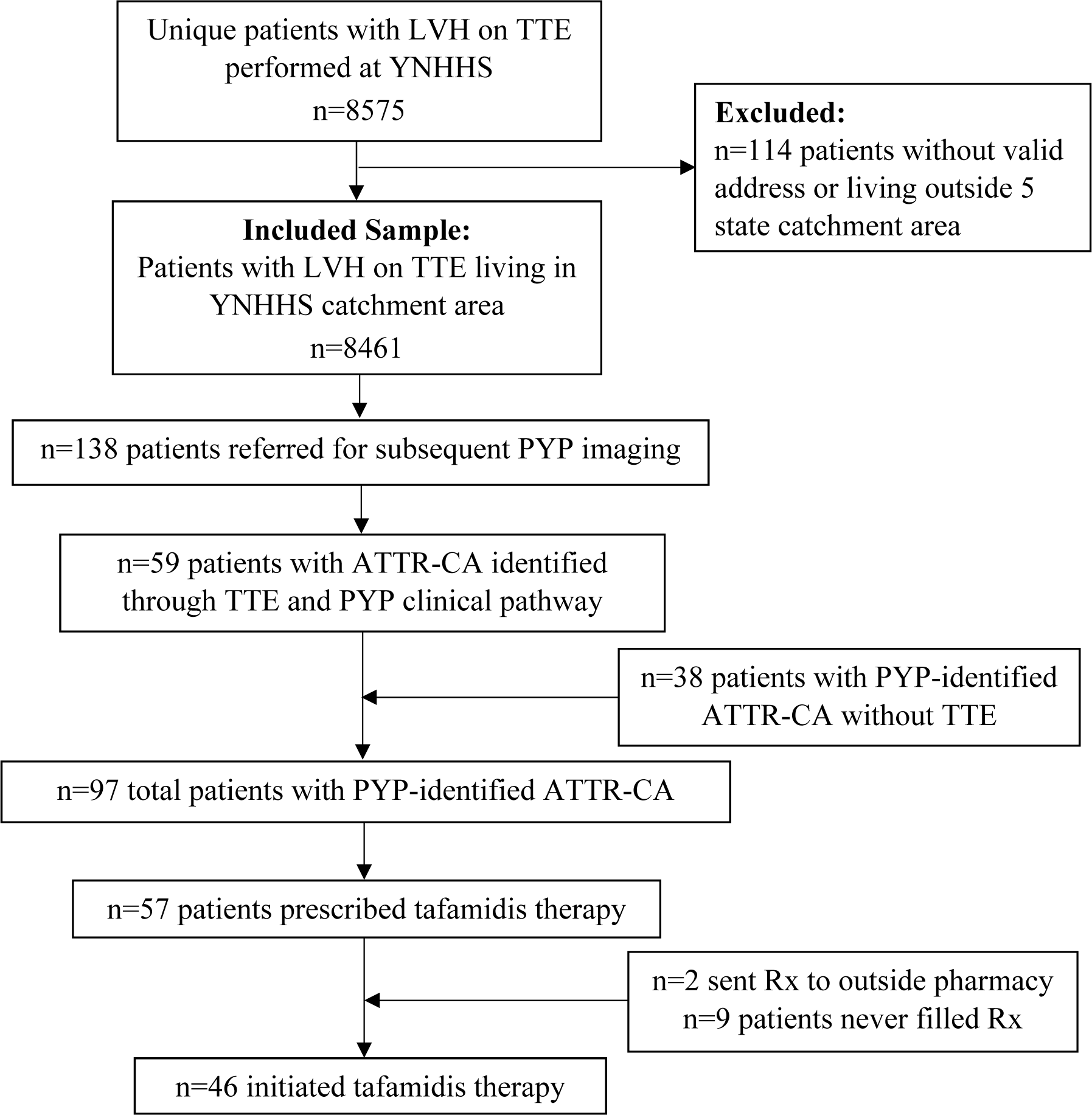
Study CONSORT diagram. TTE=transthoracic echocardiogram, YNHHS=Yale New Haven Healthcare System, LVH=left ventricular hypertrophy, PYP= technetium-99m pyrophosphate scintigraphy

### LVH and Referral to PYP

On average, patients with LVH on TTE were 69.5 (SD=13.6) years old, primarily male (70.3%), White race (76.8%), and living in areas with a low to moderate national and state ADI rank (see Table 1). Those referred for subsequent PYP imaging were older (*d*=0.45) and more likely male (*d*=-0.21) compared to patients without PYP referral. The distribution of racial categories also differed among those with vs. without PYP (*d*=-0.40), with higher PYP referral rates among patients identifying as Black/African American and American Indian/Alaskan Native, and lower rates among patients of Asian and White race (all *d*’s≥0.20). National and state ADI rank did not differ by PYP referral status (both *d*’s<0.20).

**Table 1.** Socio-demographics of patients with TTE-identified LVH by PYP completion status.

| Variable | TTE-Identified LVH<br>(n=8461) | PYP Completed<br>(n=138) | No PYP Completed<br>(n=8323) | SD |
| --- | --- | --- | --- | --- |
| Age, years | 69.5 ± 13.6 | 75.5 ± 10.5 | 69.4 ± 13.6 | **0.45** |
| Female sex | 29.7 | 22.5 | 29.8 | **-0.21** |
| Race |  |  |  | **-0.40** |
| American Indian/Alaskan Native | 0.3 | 1.5 | 0.3 | **0.97** |
| Asian | 1.1 | 0.7 | 1.1 | **-0.21** |
| Black/African American | 14.2 | 26.8 | 14.0 | **0.45** |
| Native Hawaiian/Other Pacific Islander | 0.1 | 0.0 | 0.1 | --- |
| White | 76.8 | 68.1 | 77.0 | **-0.25** |
| Other/Unknown | 7.5 | 2.9 | 7.6 | **-0.56** |
| National ADI Rank | 34.2 ± 20.8 | 36.1 ± 21.2 | 34.1 ± 20.8 | 0.10 |
| State ADI Rank | 4.8 ± 2.8 | 5.1 ± 2.9 | 4.8 ± 2.8 | 0.11 |

Values reflect mean ± standard deviation for continuous variables and percentage for categorical variables. TTE=transthoracic echocardiogram, LVH=left ventricular hypertrophy, PYP= technetium-99m pyrophosphate scintigraphy, ADI=area deprivation index, SD = standardized difference. SD values represent comparisons between patients with vs. without PYP completed with absolute values ≥ 0.20 bolded to indicate group differences equating to a small effect size or larger; negative values indicate lower mean or percentage among patients completing PYP; indeterminate when frequency for one or more groups equals 0.

### ATTR-CA and Tafamidis Prescription

Patients with ATTR-CA on PYP were an average of 81.8 (SD=7.6) years old, primarily male (81.4%) and White race (77.3%), and living in areas with a low to moderate national and state ADI rank (see Table 2). Patients prescribed tafamidis were significantly younger (*d*=-0.30) and less likely to be of other/unknown race (*d*=-0.44) compared to those not prescribed tafamidis. Patient sex, the proportions of patients of White and Black/African American race, and national and state ADI rank did not differ by tafamidis prescription status (all *d’*s<0.20).

**Table 2.**
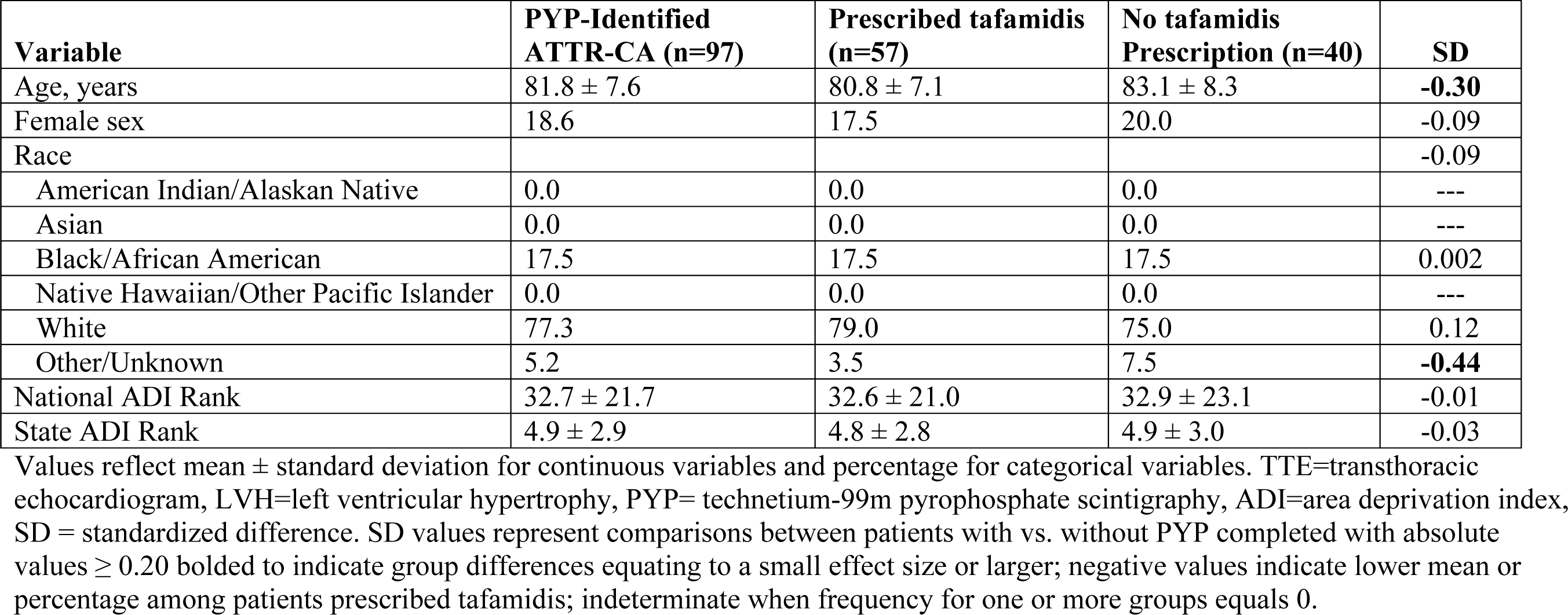
Socio-demographics of patients with PYP-Identified Cardiac ATTR-CA by tafamidis prescription status.

| Variable | PYP-Identified<br>ATTR-CA (n=97) | Prescribed tafamidis<br>(n=57) | No tafamidis<br>Prescription (n=40) | SD |
| --- | --- | --- | --- | --- |
| Age, years | 81.8 ± 7.6 | 80.8 ± 7.1 | 83.1 ± 8.3 | **-0.30** |
| Female sex | 18.6 | 17.5 | 20.0 | -0.09 |
| Race |  |  |  | -0.09 |
| American Indian/Alaskan Native | 0.0 | 0.0 | 0.0 | --- |
| Asian | 0.0 | 0.0 | 0.0 | --- |
| Black/African American | 17.5 | 17.5 | 17.5 | 0.002 |
| Native Hawaiian/Other Pacific Islander | 0.0 | 0.0 | 0.0 | --- |
| White | 77.3 | 79.0 | 75.0 | 0.12 |
| Other/Unknown | 5.2 | 3.5 | 7.5 | **-0.44** |
| National ADI Rank | 32.7 ± 21.7 | 32.6 ± 21.0 | 32.9 ± 23.1 | -0.01 |
| State ADI Rank | 4.9 ± 2.9 | 4.8 ± 2.8 | 4.9 ± 3.0 | -0.03 |

Values reflect mean ± standard deviation for continuous variables and percentage for categorical variables. TTE=transthoracic echocardiogram, LVH=left ventricular hypertrophy, PYP= technetium-99m pyrophosphate scintigraphy, ADI=area deprivation index, SD = standardized difference. SD values represent comparisons between patients with vs. without PYP completed with absolute values ≥ 0.20 bolded to indicate group differences equating to a small effect size or larger; negative values indicate lower mean or percentage among patients prescribed tafamidis; indeterminate when frequency for one or more groups equals 0.

### Tafamidis Initiation and Adherence

As detailed in Tables 2 and 3, patients with PYP-identified ATTR-CA prescribed tafamidis were, on average, 80.8 (SD=7.1) years old, primarily male (82.5%) and White race (79.0%), and living in areas with a low to moderate national and state ADI rank. Compared to patients who were prescribed tafamidis but did not initiate therapy, those who initiated therapy were older (*d*=0.64), more often of Black/African American race (*d*=0.40) and less often of other/unknown race (*d*=-0.62), and lived in areas with a higher (i.e., more economically deprived) national and state ADI rank (*d*=0.30 and *d*=0.36, respectively). Additionally, all female patients prescribed tafamidis (n=10) initiated therapy.

**Table 3.** Socio-demographics of patients with PYP-Identified ATTR-CA prescribed tafamidis by tafamidis initiation status.

| Variable | Prescribed tafamidis (n=57) | Initiated Therapy (n=46) | No Therapy Initiated (n=9)* | SD |
| --- | --- | --- | --- | --- |
| Age, years | 80.8 ± 7.1 | 81.5 ± 6.5 | 77.0 ± 9.7 | **0.64** |
| Female sex | 17.5 | 21.7 | 0.0 | --- |
| Race |  |  |  | **-0.40** |
| American Indian/Alaskan Native | 0.0 | 0.0 | 0.0 | --- |
| Asian | 0.0 | 0.0 | 0.0 | --- |
| Black/African American | 17.5 | 19.6 | 11.1 | **0.40** |
| Native Hawaiian/Other Pacific Islander | 0.0 | 0.0 | 0.0 | --- |
| White | 79.0 | 78.3 | 77.8 | 0.02 |
| Other/Unknown | 3.5 | 2.2 | 11.1 | **-0.89** |
| National ADI Rank | 32.6 ± 21.0 | 34.1 ± 20.8 | 27.8 ± 23.6 | **0.30** |
| State ADI Rank | 4.8 ± 2.8 | 5.1 ± 2.8 | 4.1 ± 2.9 | **0.36** |

Values reflect mean ± standard deviation for continuous variables and percentage for categorical variables. TTE=transthoracic echocardiogram, LVH=left ventricular hypertrophy, PYP= technetium-99m pyrophosphate scintigraphy, ADI=area deprivation index, SD = standardized difference. SD values represent comparisons between patients with vs. without PYP completed with absolute values ≥ 0.20 bolded to indicate group differences equating to a small effect size or larger; negative values indicate lower mean or percentage among patients who initiated tafamidis; indeterminate when frequency for one or more groups equals 0. \*Excludes n=2 patients who transferred their prescriptions to outside pharmacies and for whom fill records are not available.

The PDC was available for 93.5% (n=43) of the 46 patients initiating tafamidis therapy. Three patients completed only a single fill of tafamidis, thus precluding determination of this metric. Patient PDC ranged from 41% to 100% with an average of 94.2% (SD=11.0%). The threshold of PDC >80% indicating high adherence (34) was surpassed by 88.4% (n=38) of patients. PDC was not significantly associated with patient age (*r*=.06, *p*=.69), national ADI rank (*r*=.28, *p*=.07), nor state ADI rank (*r*=.25, *p*=.10). PDC also did not significantly differ by patient sex (*p*=.76) or race (*p*=.53).

## Discussion

This study is one of the first descriptions of the relationship of patient socio-demographic characteristics to an imaging-based clinical evaluation pathway for ATTR-CA, diagnosis, and referral and adherence to tafamidis therapy. These results add to our knowledge regarding ATTR-CA screening and therapeutics. First, it shows the dramatic under screening of patients with LVH for ATTR-CA in that only 1.6% of patients with LVH were referred for PYP imaging. Given estimates that suggest that between 15 to 20% of all patients with congestive heart failure and LVH have ATTR-CA (5,6), this highlights the tremendous opportunity that exists for more robust ATTR-CA screening programs. Second, on a more positive note, our data demonstrate a nearly 60% rate of tafamidis prescription in those patients ultimately identified as having ATTR-CA, and the ability of a specialty pharmacy to ensure that over 80% of those prescribed tafamidis initiated therapy. More importantly, our data demonstrate that with specialty pharmacy support there was an increase and enrichment of treatment in socio-demographic groups traditionally undertreated in cardiovascular medicine (22-24), with higher rates of tafamidis initiation among patients that are women, Black/African American, and/or living in more economically disadvantaged areas.

Of the three steps we examined along the screening to treatment clinical pathway for ATTR-CA, patient socio-demographics were most closely associated with patient drop off at the initial step from screening for LVH on TTE to subsequent PYP referral. Lower rates of PYP referral were seen among younger patients, women, and patients of Asian or White race. While these disparities are likely driven by provider knowledge of increased risk for ATTR-CA among the elderly, men, and patients of Black/African American race, strict adherence to these parameters is likely to result in the insufficient capture of patients with ATTR-CA. Indeed, among patients with hereditary ATTR-CA age of onset varies widely and may present in patients as young as 30 years old (1). In addition, wild-type ATTR-CA may occur in patients of all races, suggesting a need for a broad screening strategy that is less focused on race. While it is possible that the use of artificial intelligence algorithms and/or electronic health record prompts may enhance a the ability to enrich screening populations for patients more likely to have ATTR-CA, our sample represents real world practice at present.

Despite a relationship of TTE to PYP referral for age, sex, and race, we found a very striking lack of relationship of economic disadvantage to imaging and more importantly to tafamidis prescription and initiation. This builds on data suggesting the economics of the ATTR-CA diagnosis to treatment pathway are complex. While PYP imaging is relatively inexpensive and is routinely covered by Medicare and by most insurances, tafamidis is associated with very high costs (36). Our data suggest that a patient’s economic background as measured by the ADI did not appear to influence physicians’ decision to prescribe tafamidis and was actually associated with improved rates of therapy initiation. This suggests the possibility of a U-shaped income-to-initiation and adherence relationship with tafamidis: Patients with lower socio-economic backgrounds may have more access to Medicaid, financial assistance grants, and/or lower insurance co-pays. This may be particularly important in avoiding the Medicare ‘donut hole’ where patients are faced with paying 25% of the cost of the medication (tens of thousands of dollars per year in the case of tafamidis). Comparatively, those at the extreme high end of the income spectrum may be able to afford those costs out-of-pocket. This possibly leaves a broad class of middle-income patients less able to afford tafamidis. The complex relationship between tafamidis access, costs, and patients’ economic status is an important area of future study.

It is important to acknowledge the key beneficial role of specialty pharmacy programs in navigating access to tafamidis. These programs provide expertise in navigating benefits verification, pre-authorization, and co-pay grants programs. Our data shows that 81% of patients prescribed tafamidis initiated the medication and that 88% of patients had a high level of adherence, regardless of socio-demographic status. In comparison, rates of guideline-directed medical therapy for heart failure have been reported to the be as low as 41% (37). Thus, the present data demonstrate the powerful impact that can be obtained with comprehensive specialty pharmacy programs in driving a high level of access and adherence to tafamidis.

Findings from the present study should be interpreted within the context of the following limitations. First, although our sample represented a diversity of socio-demographic characteristics, the data were derived from a single healthcare system which may limit generalizability. Additionally, the present analyses examined the two-year period during which tafamidis first became available in the United States. It is possible that in more recent years greater provider knowledge of ATTR-CA and available therapeutics has increased PYP referral and tafamidis prescription rates. Further, our study used LVH on TTE as a potential marker for the need for ATTR-CA screening, acknowledging that incorporating other echocardiogram findings (abnormal strain, diastolic dysfunction, etc.) alone or in combination with other clinical ‘red flags’ (congestive heart failure, bilateral carpal tunnel syndrome, aortic stenosis, spinal stenosis, neuropathy, etc.) would be more likely to provide a clearer denominator of patients at risk for ATTR-CA and more likely to benefit from PYP testing. Future studies should aim to further characterize TTE to PYP referral rates for patient subgroups based on LVH etiology and the presence of other ATTR-CA risk markers. Finally, the ADI measure of socio-economics is a proxy for economic disadvantage based on patient zip code and census tract and thus may not directly reflect an individual’s true socio-economic status or access to resources.

## Conclusions

This study described the progression of patients along the multi-level clinical imaging and treatment pathway for ATTR-CA from identification of LVH on TTE to initiation of tafamidis therapy. The findings suggest potential patient subgroups that should be targeted to reduce disparities, especially in the drop off from TTE to PYP clinical imaging. Future quality improvement efforts should aim to address the tremendous opportunity for more robust ATTR-CA screening and clinical programs and further support specialty pharmacy efforts given the demonstrated benefit in ensuring the equitable provision of tafamidis therapy.

## Data Availability

N/A

